# Matrix Stiffness Influences Drug Resistance to Gemcitabine Analog and AZD 1775 Combination in PDAC Organoids

**DOI:** 10.1101/2025.06.03.25328936

**Authors:** Jonathan Barajas, Zhi Yang, Edward Agyare, Xueyou Zhu, Saun-Joo Yoon, Bo Han

## Abstract

**Background:** Pancreatic ductal adenocarcinoma (PDAC) is among the most lethal cancers, with limited therapeutic advancements and rapid development of treatment resistance. Fatty acid-conjugated gemcitabine analogs have shown improved antitumor efficacy. This study investigates the effects of gemcitabine conjugated with caprylic acid (Gemcitabine-8C) in combination with AZD 1775, a WEE1 inhibitor, using patient-derived PDAC organoids.

**Methods:** Patient-derived PDAC cells (G43, G46) were cultured in a gelatin-based 3D organoid system with tunable stiffness to mimic the tumor microenvironment. Cells were treated with gemcitabine, Gemcitabine-8C, AZD 1775, or their combination. The study assessed treatment efficacy, extracellular matrix influence, morphology, gene expression, and drug resistance mechanisms.

**Results:** The combination of AZD 1775 and Gemcitabine-8C significantly enhanced treatment efficacy compared to monotherapies or gemcitabine with AZD 1775. G43 cells were more sensitive to treatment than G46. Increased matrix stiffness correlated with greater drug resistance. Resistant cells exhibited elevated oxidative stress, while sensitive cells showed F-actin structural alterations absent in resistant counterparts.

**Conclusions:** AZD 1775 enhances the efficacy of Gemcitabine-8C at non-toxic doses, demonstrating its potential for overcoming PDAC treatment resistance. The cell origin and tumor microenvironment plays a key role in modulating drug response, highlighting the need for microenvironment and individualized-targeted strategies.

## Introduction

Pancreatic ductal adenocarcinoma (PDAC) is the most common and aggressive form of pancreatic cancer, accounting for over 90% of all pancreatic malignancies. Despite advancements in cancer research and treatment, PDAC remains a devastating disease with a dismal prognosis. The current estimated five-year survival rate is only 13%, and due to its aggressive nature and late-stage diagnosis, PDAC is projected to become the third leading cause of cancer-related death in the United States by 2030 [1, 2]

The standard-of-care treatment for PDAC often includes gemcitabine, a nucleoside analog that inhibits DNA synthesis, leading to cell cycle arrest and apoptosis. However, a major challenge in PDAC therapy is the rapid development of resistance to gemcitabine, limiting its long-term efficacy[3]. Multiple mechanisms contribute to this resistance, including altered drug metabolism, enhanced DNA repair mechanisms, and activation of survival signaling pathways[3, 4]. Given these limitations, novel therapeutic strategies that enhance gemcitabine efficacy or bypass resistance mechanisms are urgently needed.

One promising approach involves modifying gemcitabine through conjugation with fatty acids such as stearic acid, which has been shown to improve drug stability, cellular uptake, and overall therapeutic effectiveness[5]. Additionally, targeting key cell cycle regulators such as WEE1 kinase has emerged as a potential strategy to sensitize PDAC cells to DNA-damaging agents[6]. WEE1 is a crucial regulator of the G2/M checkpoint, preventing premature mitotic entry in the presence of DNA damage. Inhibiting WEE1 with small-molecule inhibitors like AZD1775 forces cancer cells to enter mitosis with unresolved DNA damage, leading to mitotic catastrophe and cell death[7]. Recent studies have demonstrated that combining AZD1775 with DNA-damaging agents can enhance therapeutic efficacy in various cancer models, including PDAC[8–10].

In this study, we investigated the therapeutic potential of AZD1775 in combination with a novel gemcitabine derivative conjugated to caprylic acid, a medium-chain fatty acid. We hypothesized that this combination would enhance cytotoxicity in PDAC cells by simultaneously improving gemcitabine bioavailability and disrupting WEE1-mediated cell cycle regulation. Our results demonstrate that the addition of AZD1775 significantly increases treatment efficacy at doses that exhibit minimal impact when used as monotherapy. This study provides insight into the potential of WEE1 inhibition as a strategy to enhance gemcitabine-based therapies in PDAC.

## Material and Methods

### Patient Derived Cells and Cell Cultures

Patient derived pancreatic adenocarcinoma cells were collected via surgical resection from the University of Florida as described previously from either Black (G43), or White (G46) Americans[11]. Cells were cultured in 10cm^2^ tissue culture dishes (Genesee Scientific, CA) in high glucose Dulbecco’s Modified Eagle’s Medium (Genesee Scientific, CA) supplemented with 10% Fetal Bovine Serum (Sigma, USA) and 1% Penicillin-Streptomycin (Genesee Scientific, CA). All cells were cultured in a humidified incubator at 37℃ and 5% CO_2_.

### Patient Derived Organoids (PDOs)

Organoids were used to study effects of Extracellular Matrix (ECM) stiffness on PDAC cells and their responses to treatment. Col-Tgel was prepared as mentioned in Fang et al.[12]. Briefly, porcine skin gelatin Type A 300 Bloom (Sigma Aldrich, MO), was dissolved in double-distilled water (ddH_2_O), sterilized and diluted in 3% or 6% final concentrations. Transglutaminase was derived from microbial source, Streptomyces mobaraensis (Ajinomoto, Tokyo, Japan). Transglutaminase was purified using SP Sepharose Fast Flow Beads (Sigma Aldrich, MO). Protein concentration of transglutaminase was determined Bradford Assay using Bio-Rad Protein Assay Dye (Hercules, CA)[13]. Cells were cultured as above then resuspended in either 3A (soft) or 6A (stiff) gel with transglutaminase mixed into the gel at a ratio of 1:20. Organoids were allowed to solidify in 5% CO_2_ at 37℃ in a humidified incubator for 45 minutes. Cultures were then grown in 10% FBS DMEM, 1% streptomycin. Cell medium was exchanged every 3-4 days for long-term cultures.

### Pharmacological Compounds

Gemcitabine was obtained from AK Scientific (Union City, CA) and Gemcitabine-8C conjugation was prepared by the Florida A&M University. AZD 1775, was purchased from MedChemExpress (Monmouth Junction, NJ). All compounds were dissolved in 90% Acetone-10% DMSO solution for stock concentrations of 10mM. Stock was then diluted in 1X PBS to working concentrations.

### Cell Cytotoxicity Assay for 2D Cultures

Cells were grown in culture as previously described, counted and seeded in 96-well tissue culture plate at a density of 1×10^4^ cells/well. Cells were then treated with gemcitabine, or it’s conjugate at 1.25µM with or without AZD 1775 at 250nM for 72 hours. Cell viability was measured using MTT Assay with absorbance read at 560nm.

### CellTiter Glo Assay for 3D Cultures

Cells were resuspended in either 3A or 6A gelatin-Tg matrices at a concentration of 1×10^6^ cells/ml. Twenty microliters (20µl) were then cast per well in a 48-well tissue culture plate (Genesee Scientific, CA) and allowed to polymerize for 45minutes at 37℃. Organoids were allowed to form for either 24 or 168 hours with medium change every 3 days for long term cultures. Both were then treated for 48 hours with 5µM gemcitabine (and analog) with or without AZD 1775 500nm combinations. To measure cell viability in PDOs, CellTiter-Glo®3D Luminescent Cell Viability Assay from Promega (Madison, Wi), was used. CellTiter-Glo was added at a ratio of 1:2 to existing medium in wells. Cultures were incubated for thirty minutes at room temperature and luminescence was read after 30 minutes.

### PCR

To examine intrinsic differences between G43 and G46 cells PCR of NRF-2 and HIF-1 was conducted. Cells were seeded in 6-well tissue culture plates at a density of 3×10^4^ cells per well. Cells were allowed to proliferate for 24 hours before RNA was extracted. Reverse-Transcription PCR (RT-PCR) was conducted followed by Real-Time PCR (qPCR) to quantify expression of ABCC1, SOX9, HIF-1. β-Actin was used a positive control. Primer sequences are listed below:

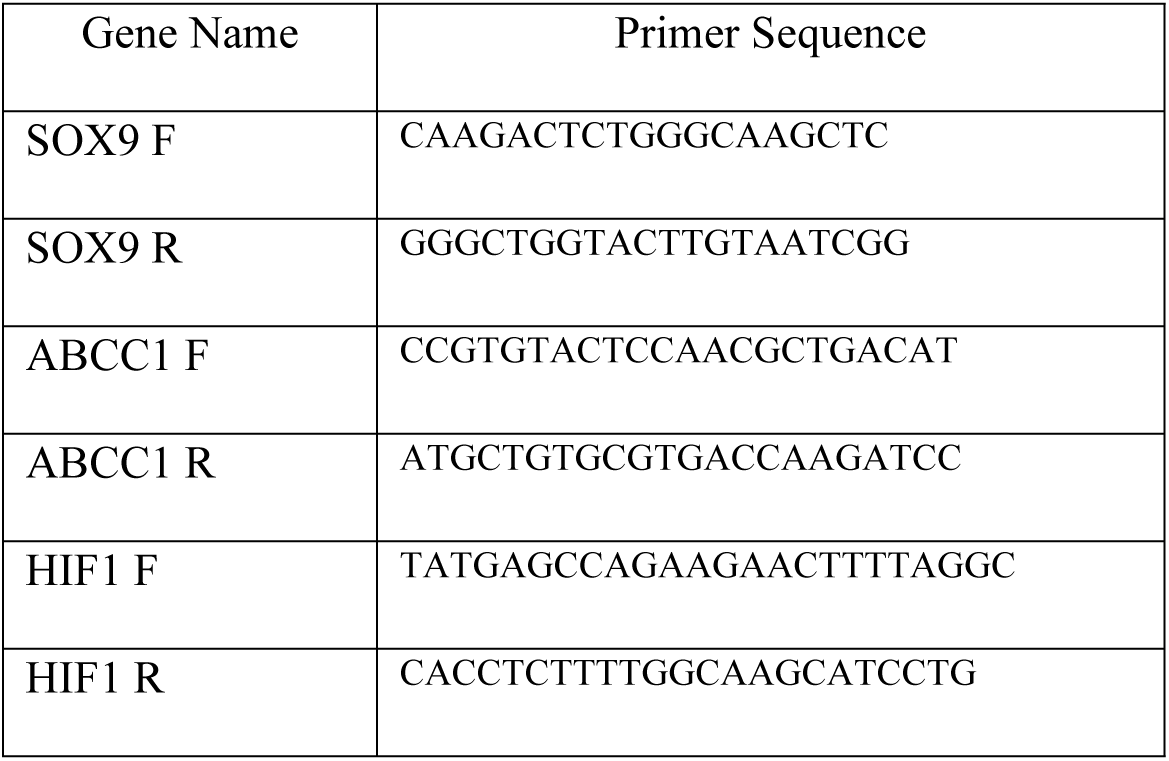

### Reactive Oxygen Species (ROS), CellROX Green Assay

Cells were plated in 6-well tissue culture plates (Genesee Scientific, CA) at a density of 3×10^4^ cells per well. Cells were allowed to incubate for 24 hours before being treated with 5µM Gemcitabine 8C and with or without 250nM AZD 1775. Cells were treated for 24 hours. After elapsed time, cells were stained with CellROX Green (Invitrogen, Carlsbad, CA), and nuclei were counterstained with DAPI dihydrochloride (Biotium Inc., CA) at a final concentration of 5µM and 1:1000 respectively in 1X PBS. Cells were cultured at 37℃ for 30 minutes. Images were captured using fluorescence microscope (EVOS, Thermo Fisher Scientific, MA).

### Phalloidin Staining

Cells were seeded and treated as described before. Fixation was achieved using Formalin for 30 minutes at room temperature. Formalin was removed by washing with 1X PBS three times. Rhodamine Phalloidin (Invitrogen, Carlsbad, CA) was prepared at a 1:10 ratio in 1% BSA and 100μl added to each well. Counterstain of nuclei was achieved using DAPI dihydrochloride (Biotium Inc., CA). Cells were incubated for 20 minutes at room temperature in the dark. Phalloidin and DAPI were removed by washing three times with 1X PBS. Cells were imaged using fluorescence microscope (EVOS, Thermo Fisher Scientific, MA).

### Live/Dead Staining

Cells were seeded in 3A or 6A gel and treated as described above in 96-well plates (Genesee Scientific, CA) and stained for Live/Dead Staining by preparing a working stock solution of 2μl Calcein-AM (Thermo Fisher Scientific, MA), 10μl EthD-1 (Thermo Fisher Scientific, MA), 5μl Hoechst 33342 (Thermo Fisher Scientific, MA), added to 10ml 1X PBS. Next, 100μl of solution was added to proper wells and incubated for 30 minutes at 37℃. The solution was washed out two times with 1X PBS and imaged using fluorescence microscope (EVOS, Thermo Fisher Scientific, MA).

### Acridine Orange Staining

Cells were seeded in 3A or 6A gel and treated as described above in 96-well plates (Genesee Scientific, CA) and stained. A working stock solution was prepared by adding 2ul acridine orange (Ambion, Invitrogen, MA) stock (1mg/ml) to 2ml 1X PBS. Once prepped, 100μl was added to proper wells. Samples were incubated at room temperature for 30 minutes and imaged using fluorescence microscope (EVOS, Thermo Fisher Scientific, MA).

### Statistical Analysis

All graphs are represented by mean ± s.d. or mean ± sem. Analysis was performed using GraphPad Prism (v. 10.4). Group analysis was achieved using Three-way ANOVA with Tukey’s Post-hoc Test. P-values are indicated by asterisk in the figures as: *P < 0.05, **P < 0.001, ***P < 0.001 and ****P < 0.0001.

## Results

### Intrinsic differences and Cell Specific Responses between G43 and G46 cells

G43 and G46 cell lines, derived from different patient donors with distinct racial backgrounds, G43 from a Black male and G46 from a White female exhibit notable differences in morphology, gene expression, and functional behavior. Given the potential influence of patient-specific factors on tumor biology and therapeutic responses, characterizing these intrinsic differences provides valuable insights into the variability in treatment efficacy observed in further studies. As shown in Fig. 1A, G43 cells exhibit a more differentiated morphology characterized by a cobblestone-like, epithelial-like appearance, suggesting that these cells are more committed to an epithelial lineage. Differentiated cells typically display reduced plasticity and limited proliferative potential, which may contribute to increased sensitivity to cytotoxic agents observed in subsequent treatment studies. In contrast, G46 cells demonstrate a “honeycomb” pattern, which is commonly associated with a stem-like or progenitor state. This morphology suggests a higher degree of plasticity and self-renewal capacity, consistent with the maintenance of a stem-like phenotype. Cells exhibiting this pattern often have a greater ability to survive therapeutic stress, contributing to treatment resistance. The honeycomb appearance is reminiscent of mesenchymal-like or epithelial-to-mesenchymal transition (EMT)-like morphology, which correlates with increased invasiveness, migration, and drug resistance in PDAC.

**Fig. 1.**
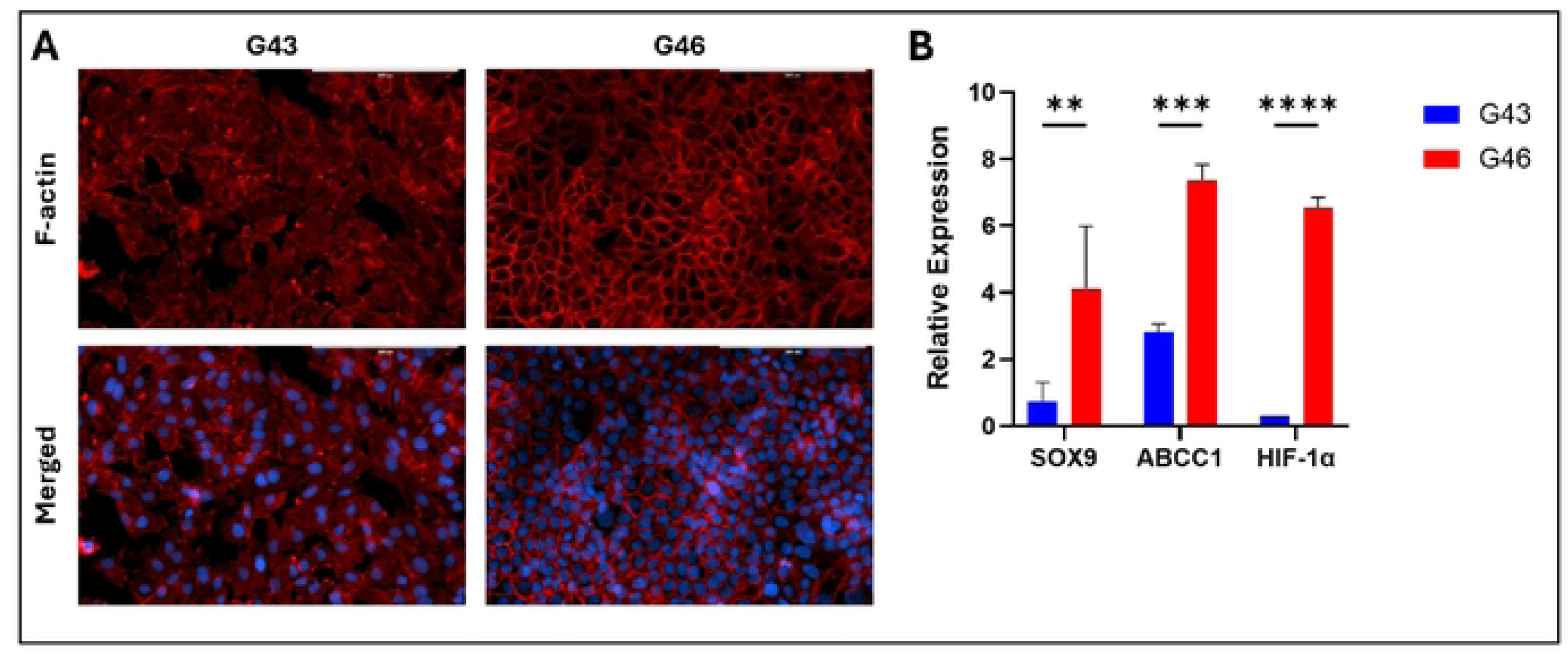
Intrinsic Differences between G43 and G4S cells. (A) F-actin staining of G43 cells and G46 cells. Red, F-actin; Blue, DAPI. Scale bars 200µM. (B) RT-PCR quantification of sternness (SOX9), drug resistance (ABCC1) and oxidative stress (HIF-1a) markers. Data represents mean ± SD from biological triplicates. **P < 0.01, ***P < 0.001 and ****P < 0.0001.

### Cell Specific Responses to Gemcitabine and Gemcitabine 8C

To further investigate the differences in treatment responses between G43 and G46 cells, both cell lines were treated with either gemcitabine or its caprylic acid-conjugated analog, Gemcitabine 8C in 2D cultures to develop a baseline response. Following treatment, cell viability was assessed using MTT Assay. To visualize possible effects of cell cycle arrest, morphology was visualized using phalloidin. Cells were stained with phalloidin to visualize F-actin, a key cytoskeletal component responsible for maintaining cell shape, structure, and motility. As anticipated, G43 cells demonstrated greater sensitivity to treatment, with a more pronounced reduction in F-actin staining upon exposure to Gemcitabine 8C compared to gemcitabine alone (Figure 2A and 2B). G43 cell viability was also lower when treated with Gemcitabine 8C as compared to gemcitabine alone (Figure 2B). In contrast, G46 cells exhibited a smaller reduction in F-actin staining following treatment with Gemcitabine 8C (Figure 2C). G46 cells also showed an expected resistance to treatment with both gemcitabine and Gemcitabine 8C at all doses (Figure 2D).

**Fig. 2.**
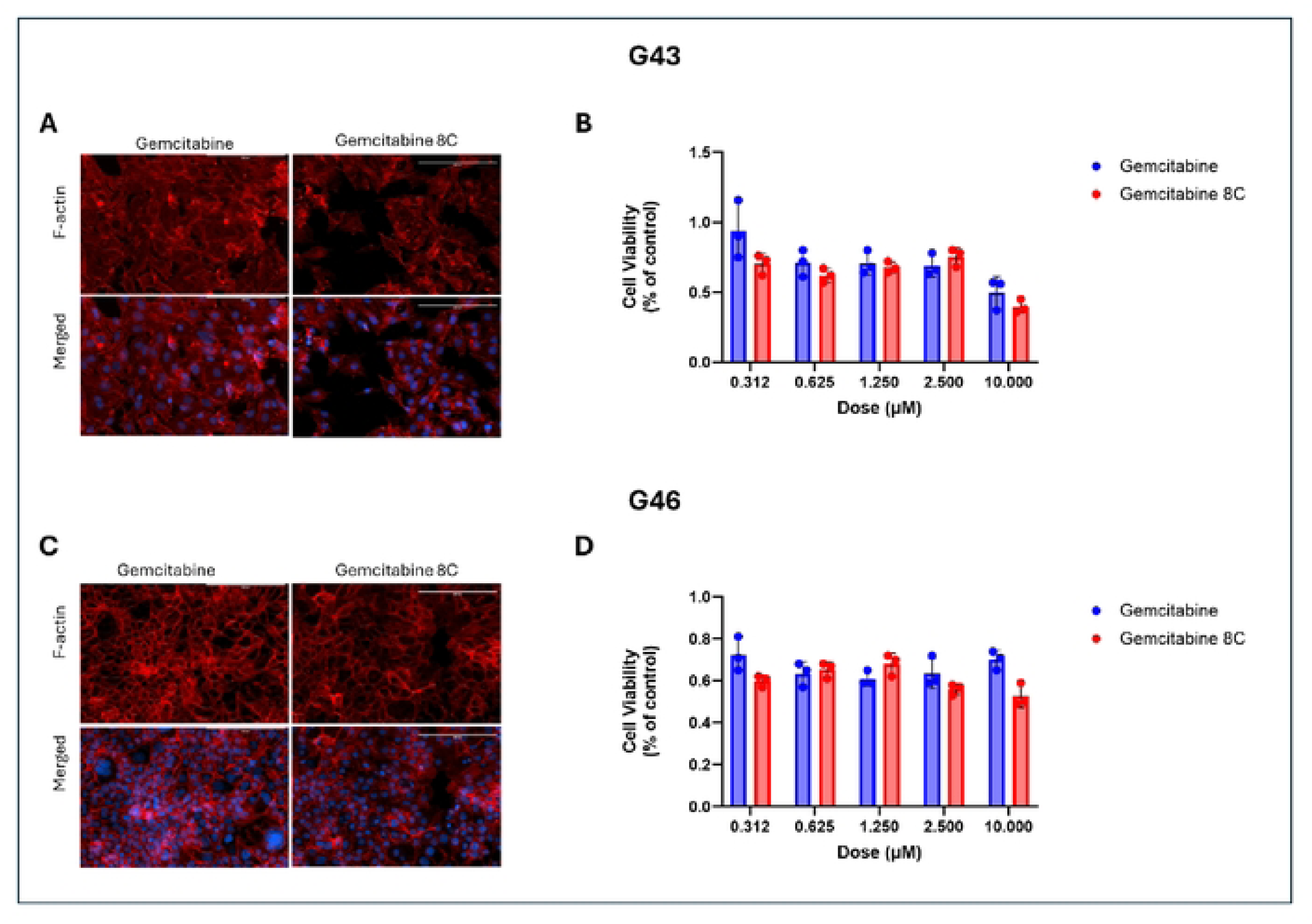
Effects of gemcitabine and gemcitabine analog treatment on sensitive and resistant cells. (A) F-actin staining of sensitive, G43, or resistant (C), G46, cells after treatment with either gemcitabine or gemcitabine BC. (8) Cell viability of (A) G43 and (D) cell viability of (C) G46 cells. Data represents means± s.d. n=3 biological replicates. Red, F-actin; Blue, DAPI. Scale bars 200µM.

### AZD 1775 in Combination with Gemcitabine or Gemcitabine Analog Increases Efficacy

To assess the effects of AZD 1775, a potent WEE1 inhibitor, on PDAC cells, G43 and G46 cells were treated with either gemcitabine or Gemcitabine 8C, alone or in combination with AZD 1775. As previously shown in Figure 2, G43 cells exhibited greater sensitivity to treatment, a trend that was further confirmed in Figure 3A and 3C. In G43 cells, treatment with Gemcitabine 8C and AZD 1775 led to a significant reduction in F-actin staining. In contrast, G46 cells did not exhibit a significant decrease in F-actin fluorescence following treatment with either gemcitabine or Gemcitabine 8C alone, as shown in Figure 3B. However, a significant difference in cell viability was observed between monotherapy and combination treatment (p < 0.01) (Figure 3D).

**Fig. 3.**
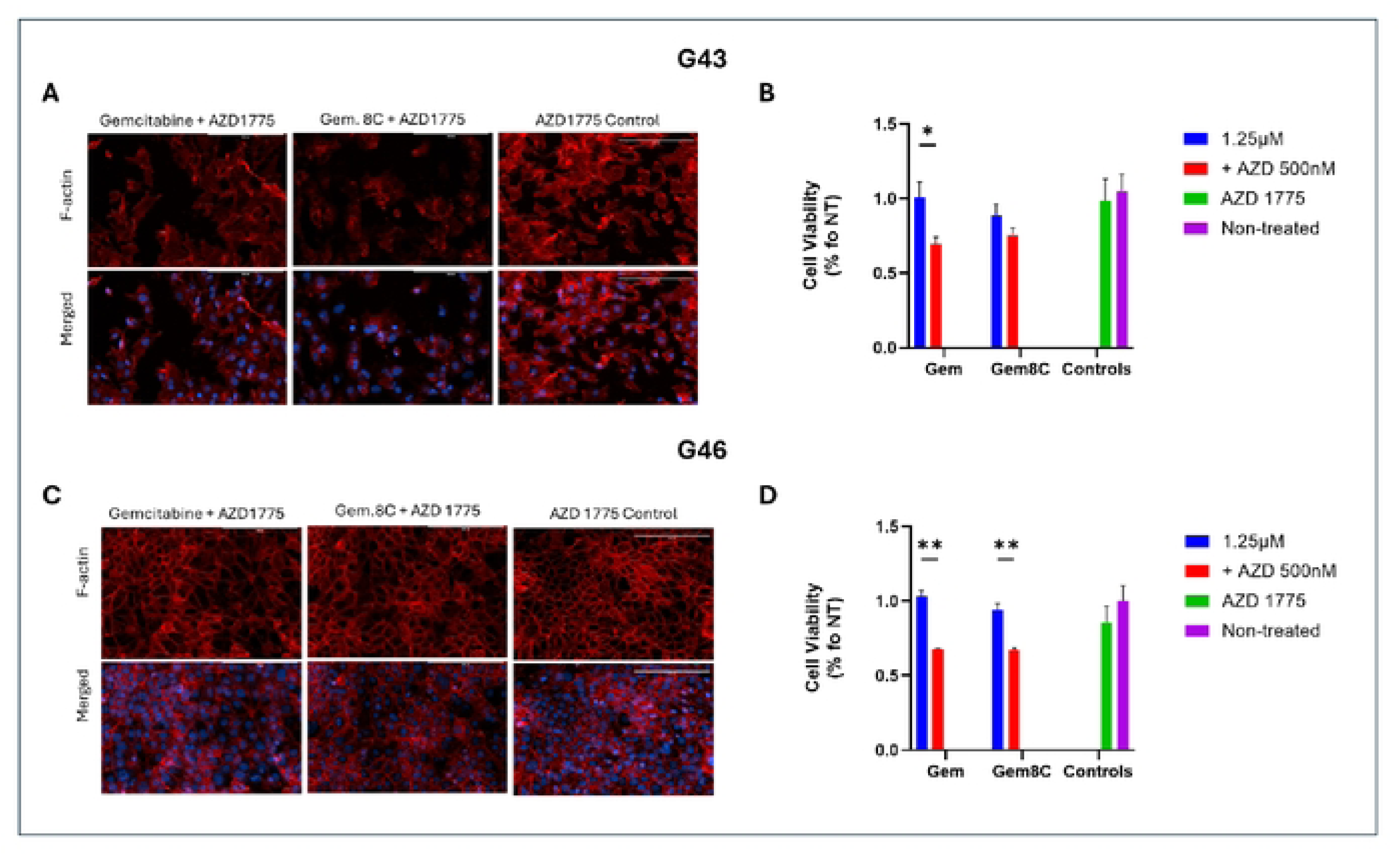
Effects of AZD 1775 combinational treatment. (A) F-actin staining of sensitive, G43, or resistant (B), G46, cells by phalloidin staining, 24hrs after treatment with either gemcitabine or Gemcitabine 8C ± AZD 1775. (C) Percent viability of G43 or (D) G46 cells after treatment. For viability assay’s, n=3 biological replicates for each group. Data represents means± s.d. Red, F-actin; Blue, DAPI. Scale bars 200µM. *P < 0.05, **P < 0.01.

### AZD 1775 Treatment increases Oxidative Stress in Resistant Cells

To examine the effect that combinational treatment with AZD 1775 produces, cells were treated with Gemcitabine 8C either in combination with or without AZD 1775. After treatment, CellROX Green was used as a probe for reactive oxygen species (ROS). Treatment with gemcitabine 8C alone did not produce significant ROS in both G43 and G46. There was a significant increase in ROS production in G46 as compared to G43 when treated in combination (p < 0.01) (Figure 4C). However, the greatest increase in ROS production was seen in G46 treated with both drugs as compared to G46 treated with gemcitabine 8C alone (p < 0.001) (Figure 4C). These finding are consistent with other findings that double-stranded DNA breaks can induce oxidative stress[14].

**Fig. 4.**
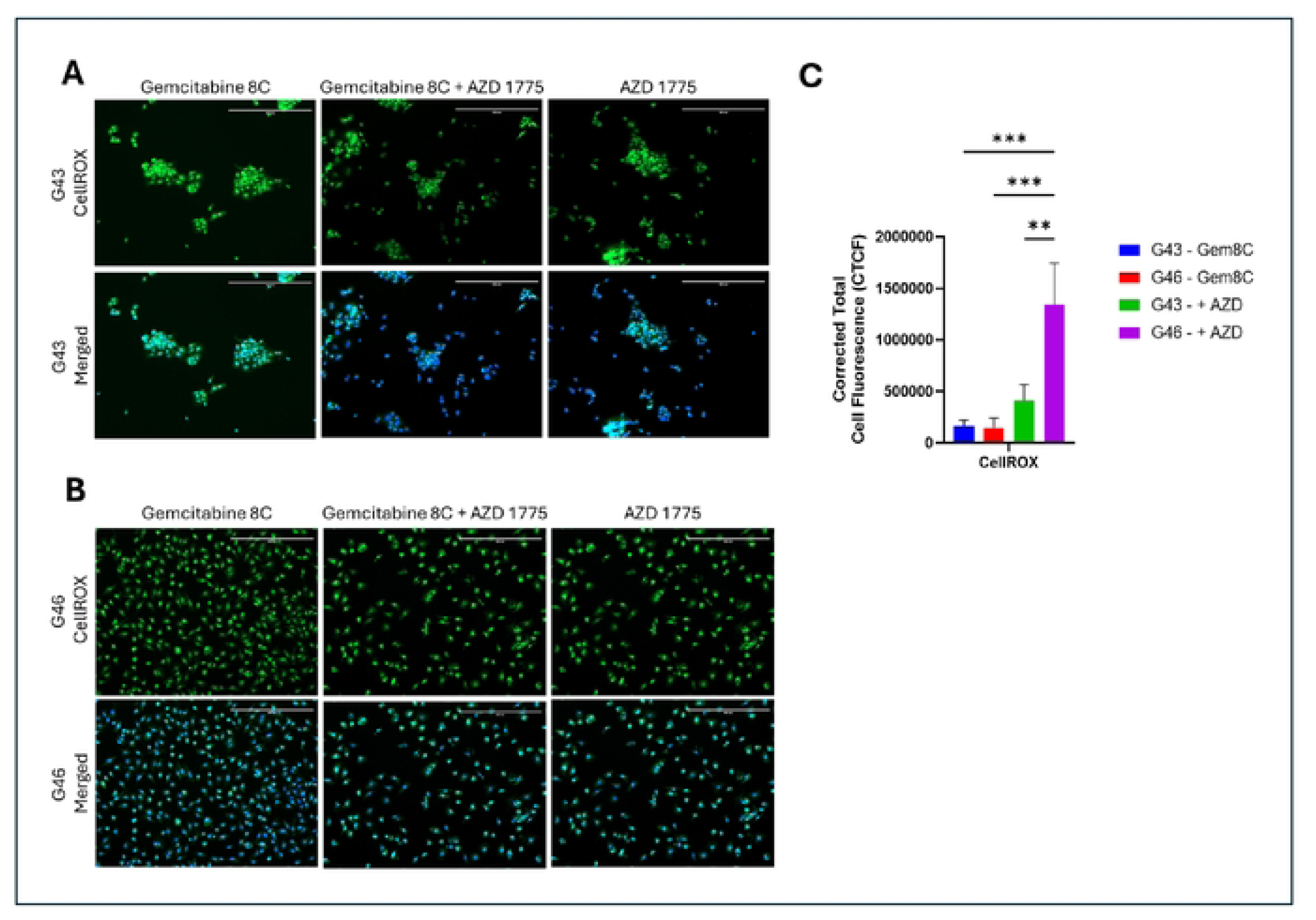
Induced oxidative stress in PDAC Cells. (A) G43 cells and (8) G46 cells were treated with Gemcitabine 8C (SµM) with or without AZD 1775 (250nM) for 24 hours. Cells were then stained for oxidative stress (CellROX) and nuclei counterstained with DAPI. (C) Quantitative representation of ROS. Images were captured using fluorescence microscope. Analysis of reactive oxygen species was done using lmageJ. Green, CellROX; Blue, DAPI. Scale bars 400µM. For all represented data (n=3) ± SD. **p < 0.01 and ***P < 0.001

### Organoid formation in varying matrix stiffness influences morphology

To investigate the impact of extracellular matrix (ECM) stiffness on tumor cell morphology and cluster formation, G43 and G46 cells were embedded in Col-Tgels with two distinct stiffness conditions: soft (3A) and stiff (6A). Tumor cells were allowed to incubate within the matrices for either 48 hours (short-term) or 168 hours (long-term) to assess the effects of matrix stiffness and incubation time on cellular and organoid development. In G43 cells, cluster formation was observed in both soft and stiff matrices at both time points; however, key differences in cluster size and organization emerged with prolonged incubation and increased matrix stiffness (Figure 5A).

**Fig. 5.**
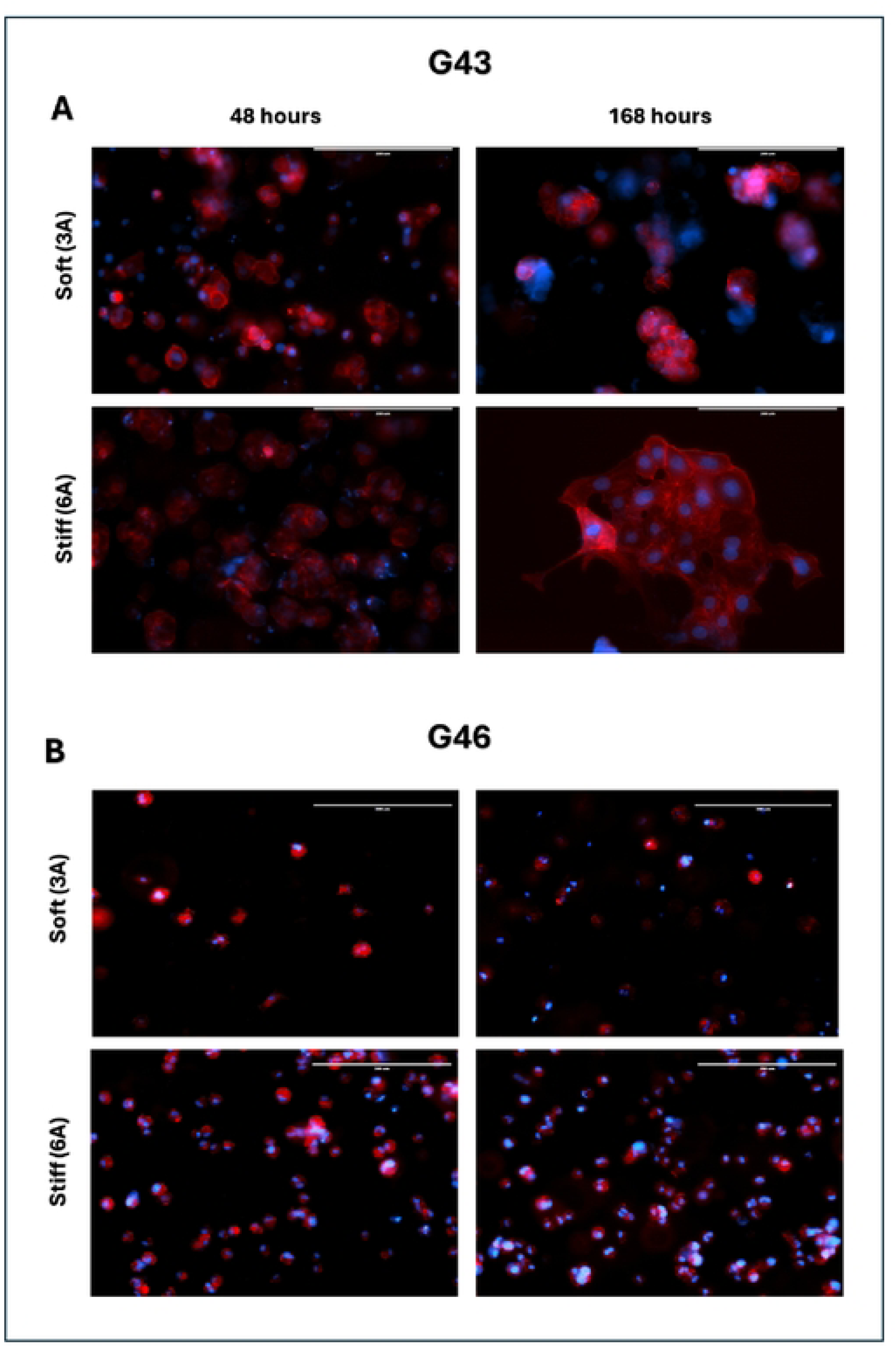
Matrix stiffness influences cluster formation over time. (A) G43 or (8) G46 cells were seeded and allowed to proliferate for either 48 or 168 hours. Organoids were then fixed with 4% formalin and F-actin was visualized using phalloidin. Nuclei were counterstained with DAPI. Images captured using fluorescence microscope. Red, phalloidin; Blue, DAPI. Magnification, 20X. Scale bars 200µM.

At 48 hours, G43 cells exhibited the early formation of small, loosely packed clusters, suggesting cell-cell adhesion and early organoid development. Cluster formation was evident in both soft and stiff matrices, although clusters appeared more compact and slightly larger in the stiff matrix. After 168 hours, G43 cells in the stiff matrix demonstrated significantly larger and denser clusters, consistent with increased proliferation, cell-cell adhesion, and maturation of organoid structures over time. The larger cluster size observed in stiffer environments suggests that ECM rigidity may promote growth and organization of tumor spheroids, by potentially providing biochemical cues that enhance cell signaling.

In contrast, G46 cells exhibited a markedly different response to matrix stiffness and incubation time, assuming a predominantly single-cell phenotype with minimal cluster formation (Figure 5B). At 48 hours, G46 cells remained largely dispersed in both soft and stiff matrices, failing to establish organized clusters. The absence of early cluster formation suggests that G46 cells may exhibit impaired cell-cell signaling compared to G43 cells. Even after prolonged incubation (168 hours), G46 cells maintained a dispersed, single-cell phenotype, with little to no evidence of cluster formation. This suggests that matrix stiffness alone was insufficient to drive cell aggregation or organoid formation in G46 cells. The results may indicate that these cells may be inherently less capable of forming structured clusters or may rely on different signaling cues for cell aggregation and organoid development.

### AZD 1775 enhances efficacy of Gemcitabine 8C in Patient-Derived Organoids (PDOs)

To explore the impact of AZD 1775 on the efficacy of gemcitabine 8C, we conducted a series of experiments in pancreatic ductal adenocarcinoma (PDO) cells seeded in 3D matrices, which mimic the extracellular matrix (ECM) stiffness commonly observed in tumor microenvironments. Cells were treated with either Gemcitabine 8C after a designated growth period (48 hours or 168 hours) or combination of Gemcitabine 8C + AZD 1775 (250nM). Cell viability was assessed using Celltiter Glo 3D Viability Assay and Live/Dead staining.

In the 48-hour organoid formation time, cells responded better to treatment in stiffer (6A) ECM matrices. Across both G43 and G46, there was a decrease in viability as determined by ATP luminescence from the CellTiter assay (Figure 6A). In the live/dead staining, for both cells, there is an increase in live staining from Gemcitabine 8C alone in stiffer matrices. However, in combination treatment, there is an increase in live staining seen in soft matrices (3A) (Figure 6B).

**Fig. 6.**
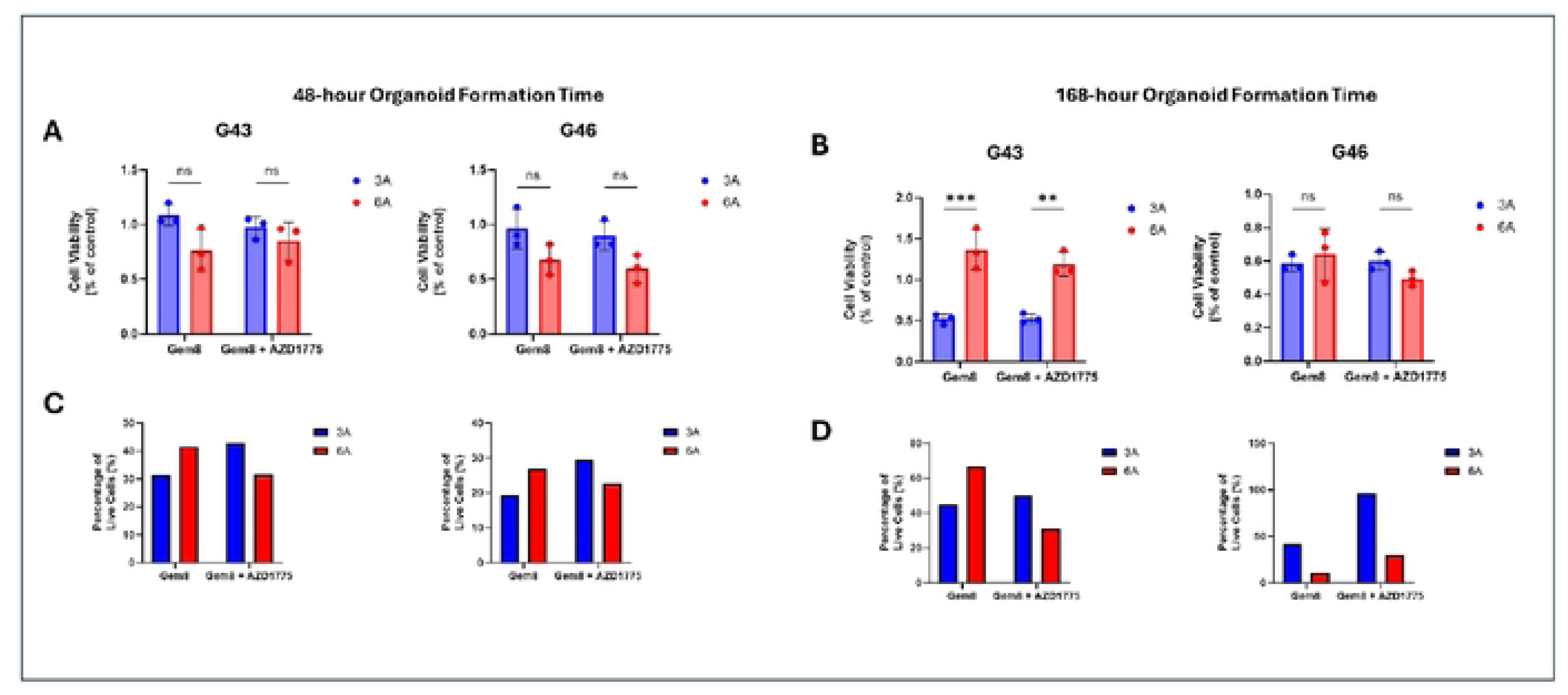
Celltiter Glo 3D viability assay after treatment with Gemcitabine 8C or in combination with AZD 1775. (A) G43 or G46 cells treated with Gemcitabine 8C (1.25µM) or Gemcitabine 8C + AZD 1775 (250nM) after 48 hour organoid culturing time in either soft (3A) or stiff (6A) ECM conditions. (B) G43 or G46 cells treated with Gemcitabine 8C (1.25µM) or Gemcitabine 8C + AZD 1775 (250nM) after 48 hour organoid culturing time in either soft (3A) or stiff (6A) ECM conditions. (C) Analysis of live/dead staining using lmageJ (v.1.54j) of conditions from (A). (D) Analysis of live/dead staining using lmageJ (v.1.54j) of conditions from (C). For all data represented, n = 3 ± SD.

When G43 cells were given time to grow in organoids, a reversal in trends was observed, viability was seen to be increased in stiff ECM (6A) in Gemcitabine 8C. However, there was a discrepancy between combination therapy where there is an increase in luminescence readings (Figure 7C) but a decrease in live/dead staining (Figure 6D). In G46 cells this trend is appreciated fully, where there is an increase in luminescence readings in both Gemcitabine 8C and Gemcitabine 8C + AZD 1775 (Figure 6C) but a drastic decrease in live/dead staining (Figure 6D).

**Fig. 7:**
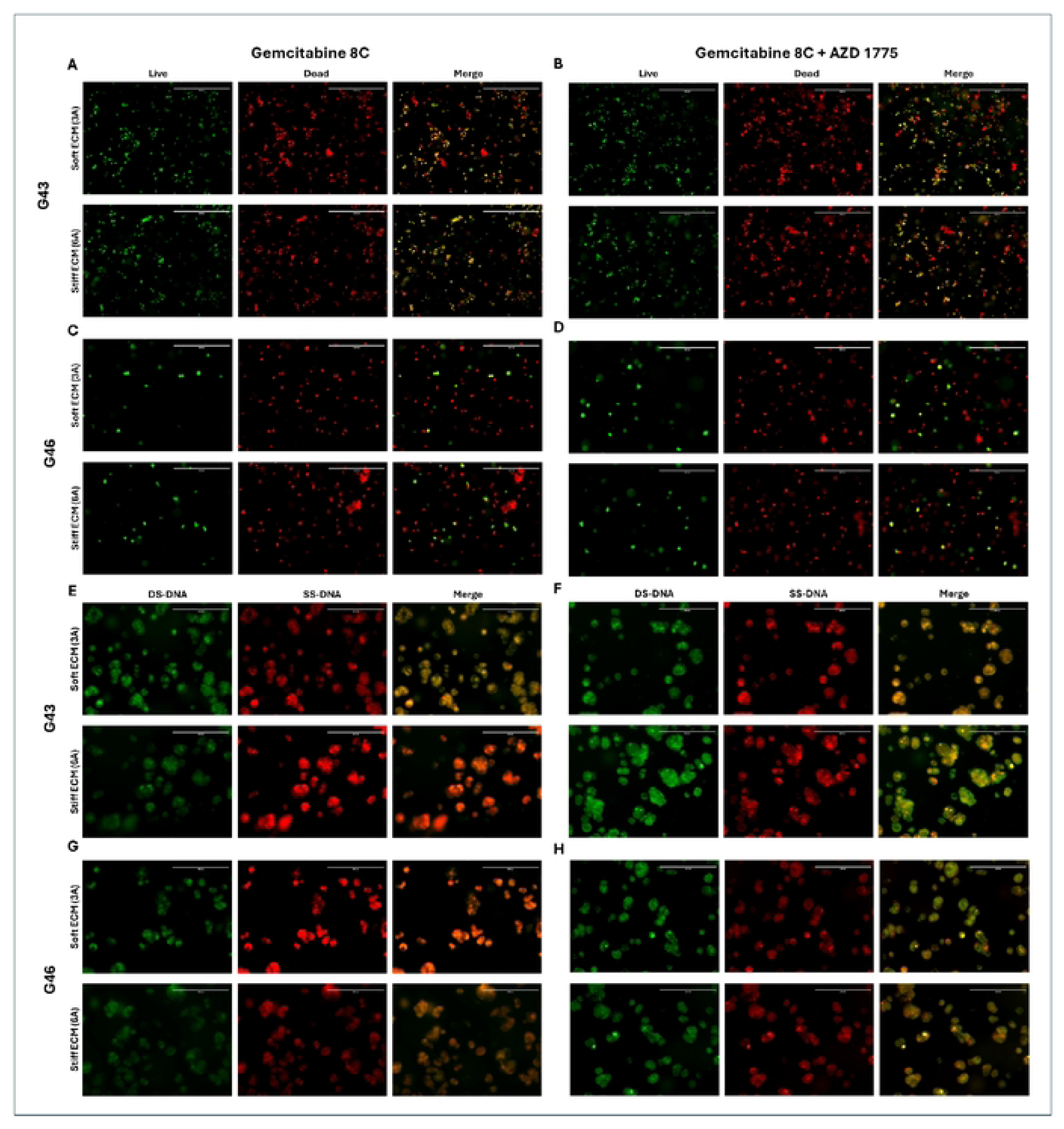
4S-hour Organoid Formation Live Dead and Acridine Orange Staining. Live/Dead labeling of (A, C) Gemcitabine SC (1.25µM) or (B, D) Gemcitabine SC + AZD 1775 (250nM) treated G43 and G46 cells, respectively. Acridine orange staining of (E, G) Gemcitabine SC or (F, H) Gemcitabine SC+ AZD 1775 treated G43 and G46 cells, respectively.

**Fig. 8:**
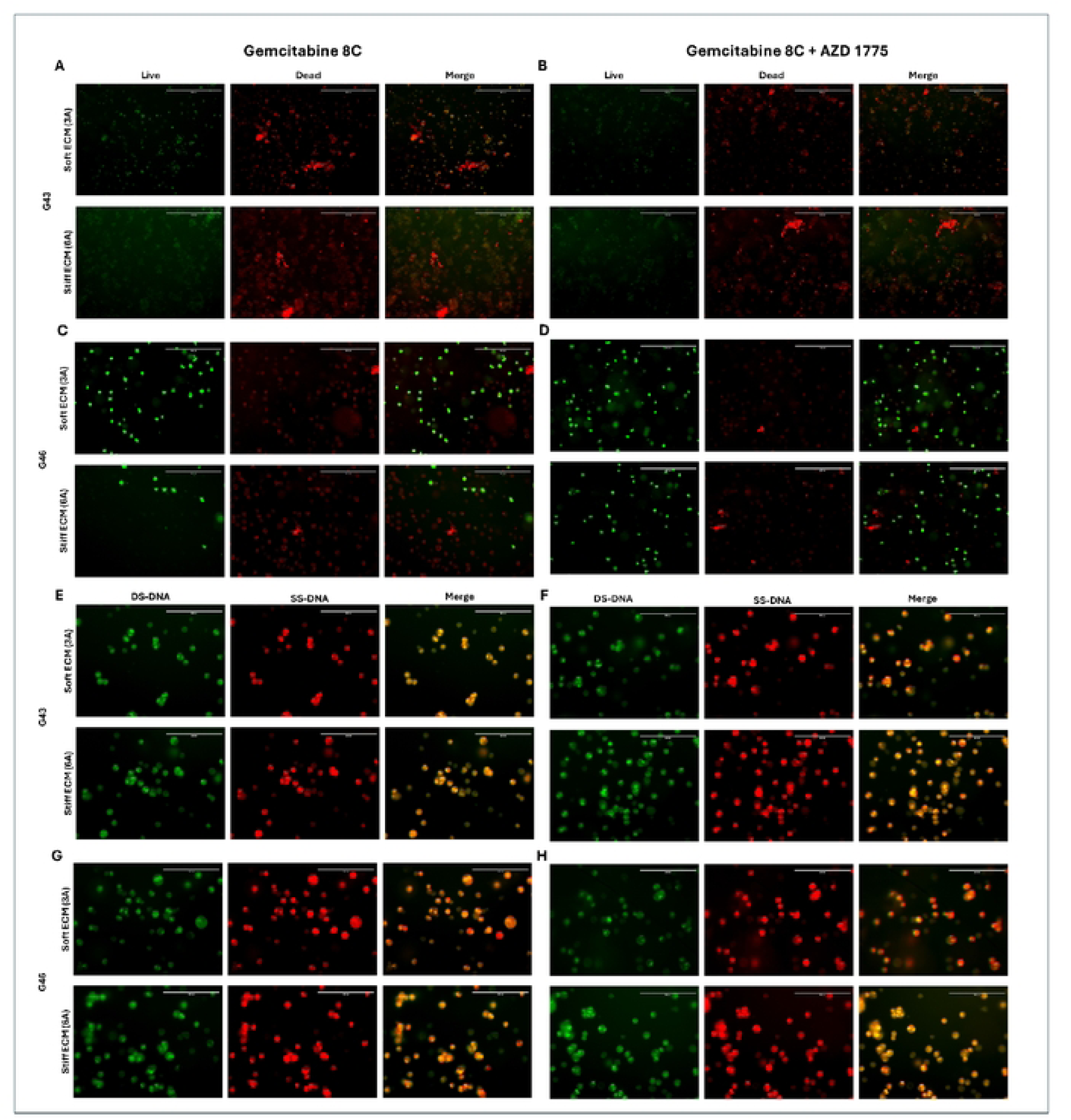
16B-hour Organoid Formation Live Dead and Acridine Orange Staining. Live/Dead labeling of (A, C) Gemcitabine BC (1 25µM) or (B, D) Gemcitabine BC + AZD 1775 (250nM) treated G43 and G46 cells, respectively. Acridine orange staining of (E, G) Gemcitabine BC or (F, H) Gemcitabine BC+ AZD 1775 treated G43 and G46 cells, respectively. Live/Dead staining; Green, live; Red, dead. Acridine Orange; Green. DS­ DNA; Red, SS-DNA. Scale bars 200µM DS-DNA (double-stranded DNA). SS-DNA (single-stranded DNA).

**Fig. 9.**
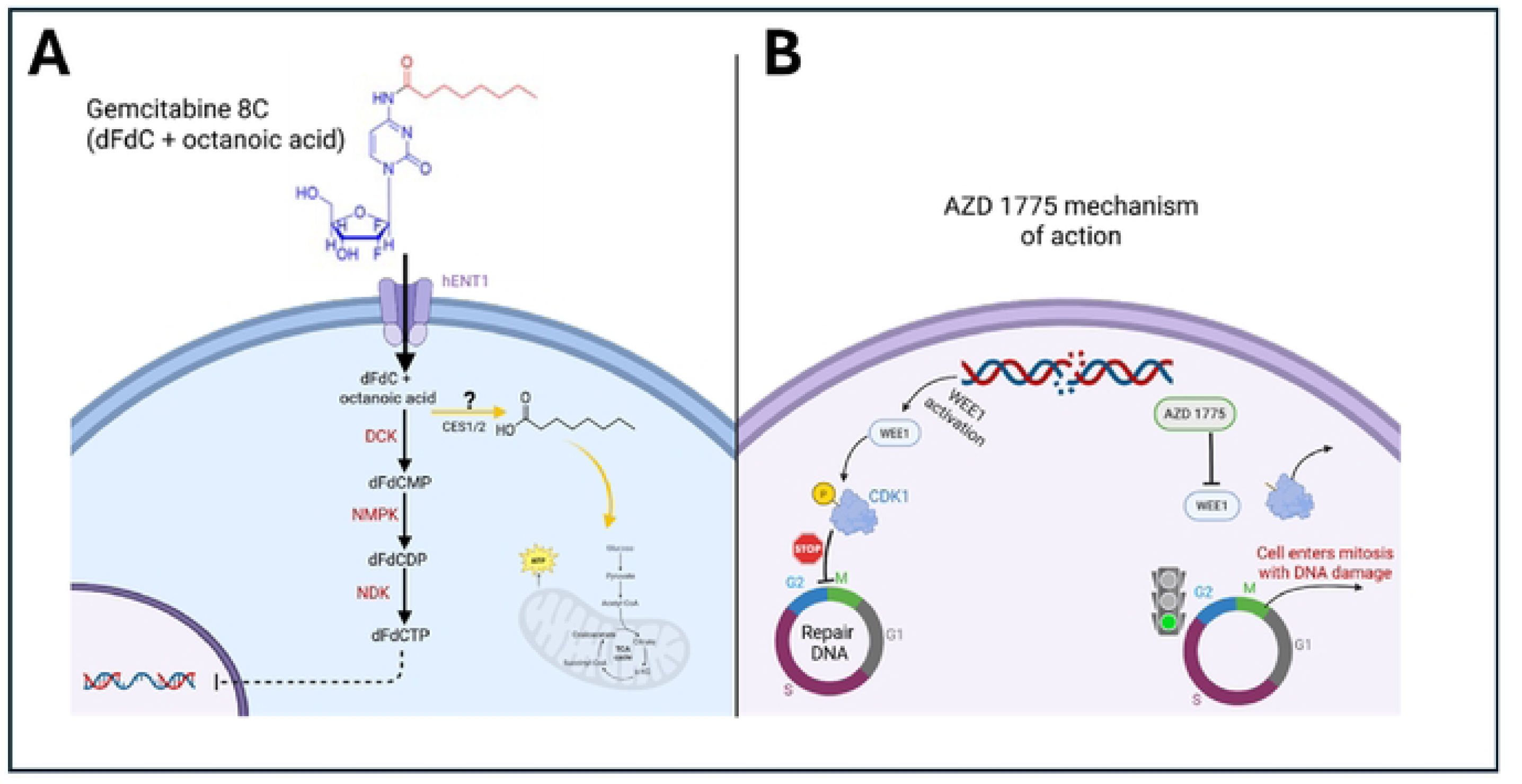
Proposed mechanism of action of Gemcitabine SC and AZD 1775. (A) Gemcitabine (dFdC, 2’,2’-difluoro-2’-deoxycytidine) enters the cell via hENT1 transporters and is phosphorylated by deoxycytidine kinase (DCK), nucleoside monophosphate kinase (NMPK), and nucleoside diphosphate kinase (NDK) to its active form, dFdCTP. dFdCTP incorporates into DNA, causing replication stress and DNA damage. We hypothesize that Gemcitabine 8C, which includes an octanoic acid moiety, may be cleaved (possibly by CES1/2) to release octanoic acid, which could feed into the TCA cycle, contributing to ATP production and anaplerosis. (B) AZD 1775 inhibits WEE1. a kinase that phosphorylates CDK1 to enforce the G2/M checkpoint. WEE1 inhibition allows cells with DNA damage to prematurely enter mitosis, resulting in mitotic catastrophe and cell death. Abbreviations: dFdCMP, dFdC monophosphate; dFdCDP, dFdC diphosphate; DCK, deoxycytidine kinase; NMPK, nucleoside monophosphate kinase; NDK, nucleoside diphosphate kinase; hENT1, human equilibrative nucleoside transporter 1; CDK1, cyclin-dependent kinase 1.

### Treatment with AZD 1775 Increases Cell Death and Inhibits Cell Cycle – 48 hour PDO Formation

The combination effects of Gemcitabine 8C + AZD 1775 were tested against G43 and G46 cells in either soft (3A) or stiff (6A) ECM gels and quantified using Live/Dead staining. To also further study the proposed mechanism of AZD 1775 acting on the cell cycle, cells were treated with Gemcitabine 8C + AZD 1775 and stained with acridine orange, with green fluorescence for double stranded DNA and red fluorescence for single stranded DNA.

In G43 cells there is an increase in cell viability seen in Gemcitabine 8C + AZD 1775 treatment in soft (3A) ECM. However, in stiff ECM (6A), there is a decrease in live cells seen in the combination treatment arm (Figure 7A and 7B). When stained for double or single stranded DNA, there is an increase in single-stranded DNA (SS-DNA) in stiff ECM (6A) when treated with Gemcitabine 8C alone (Figure 7E). However, when treated with AZD 1775, there is an increase in double-stranded DNA (DS-DNA).

In the G46 cells there is a similar trend observed. There is an increase in live cell staining in the combinational treatment in soft (3A) ECM but a decrease in stiff (6A) ECM in combinational treatment. When stained with acridine orange, there is an increase in single-stranded DNA (SS-DNA) in soft (3A) ECM in Gemcitabine 8C treated cells and an increase in double-stranded DNA (DS-DNA) in both soft and stiff ECM when treated in combination with AZD 1775 (Figure 7G and 7H).

### Treatment with AZD 1775 Increases Cell Death and Inhibits Cell Cycle – 168-hour PDO Formation

To capture any effects of long-term incubation in matrices of varying stiffnesses, cells were incubated for 168-hours (7-days) before treatment with Gemcitabine 8C or Gemcitabine 8C + AZD 1775. Viability was assessed using Live/Dead staining. For comparison studies DNA content was also assessed using acridine orange as before, with green for single-stranded DNA (SS-DNA) and red for single-stranded DNA (SS-DNA).

In the long-term culture, in G43 cells, in both soft (3A) and stiff (6A) ECM, there was increase in cell death as shown by an overall reddish coloration of the merged live/dead fluorescence in both Gemcitabine 8C alone and in combination. When treated with both treatments, there is slightly more single-stranded DNA or acidic conditions being detected by the acridine orange staining.

When G46 cells were treated with Gemcitabine 8C or in combination there was a decrease in viability in both treatment regimens in both soft (3A) or stiff (6A) matrices. After treatment, cells were stained with acridine orange, and had similar results to G43, where there is slightly more single-stranded DNA in acidic conditions as shown by an increase in a yellowish tint when images were merged.

### Treatment with Gemcitabine 8C Influences ATP-based Viability Assays

As was seen in figs 6-8, there is a discrepancy between live/dead staining and ATP readings, using the CellTiter-Glo 3D Cell Viability Assay. The results maybe explained by the structure of the gemcitabine analog. It is known from previous literature that carboxylesterases, play an important role in the metabolism of drugs. Here, it may be possible that the octanoic acid is being cleaved from the parent drug gemcitabine and is being incorporated into the TCA cycle to produce increased ATP levels. The following diagram shows a potential schematic of how this may occur.

## Discussion

This study evaluated the combinational effect of a DNA-damaging agent and a WEE1 inhibitor (AZD 1775) in patient-derived organoids (PDOs) generated from pancreatic ductal adenocarcinoma (PDAC) cells. Our results demonstrated that the combination of AZD 1775 and gemcitabine conjugated to caprylic acid (Gemcitabine 8C) was more effective than either drug alone, offering a novel therapeutic approach with potential to overcome treatment resistance. Given the limited treatment options and poor prognosis associated with PDAC, these findings are highly promising. Furthermore, PDAC remains largely unresponsive to immunotherapy due to the dense stroma and the complex tumor microenvironment (TME), making it imperative to explore alternative treatment strategies that target intrinsic vulnerabilities within PDAC cells.

Our study highlights intrinsic differences between the G43 and G46 cell lines, which contributed to distinct treatment responses. Notably, G46 cells exhibited significantly higher SOX9 expression compared to G43 cells (Figure 1B), consistent with the known role of SOX9 in maintaining stem-like characteristics and enhancing chemoresistance in PDAC cells [15, 16]. Morphological differences between the two cell types, captured in Figure 1A, further support these findings. Additionally, G46 cells demonstrated increased expression of ABCC1, a multidrug resistance-associated protein known to mediate chemoresistance by effluxing chemotherapeutic agents [17]. These observations suggest that G46 cells possess a more resistant phenotype, correlating with higher treatment resistance observed in our experiments.

Interestingly, treatment with AZD 1775 and gemcitabine 8C disrupted the F-actin cytoskeleton, particularly in the more sensitive G43 cells. Although the precise mechanism behind this disruption remains unclear, it is likely linked to the mitotic effects of AZD 1775. Under normal conditions, WEE1 phosphorylates Cyclin-Dependent Kinase 1 (CDK1) in response to DNA damage, arresting the cell cycle at the G2/M checkpoint to allow DNA repair [9]. However, when WEE1 is inhibited by AZD 1775 in the presence of DNA damage, CDK1 remains unphosphorylated, forcing the cells to prematurely enter mitosis. Given the well-established role of the Rho family of GTPases in regulating actin cytoskeleton dynamics, and their emerging role in the DNA damage response (DDR) pathway [18], it is plausible that dysregulation of GTPase activity due to DNA damage and forced mitotic entry may contribute to the observed disruption of F-actin. This opens new avenues for investigating how mitotic dysregulation and actin cytoskeleton disruption may affect treatment outcomes.

Treatment with AZD 1775 significantly increased oxidative stress in both G43 and G46 cells; however, oxidative stress was notably higher in G46 cells (Figure 4A, B). This phenomenon is consistent with evidence suggesting that double-stranded DNA (DS-DNA) breaks can induce oxidative stress, contributing to cellular damage and potentially influencing treatment outcomes [14, 19]. The heightened oxidative stress observed in G46 cells may further explain their increased resistance, possibly by activating stress-response pathways that enable survival despite genotoxic stress.

The use of patient-derived organoids (PDOs) provided an opportunity to model the complex TME in PDAC, particularly through the incorporation of collagen-based (Col-T) gels with varying stiffness. Softer ECM (3A gel) models the microenvironment of early-stage or less dense PDAC, whereas stiffer ECM (6B gel) mimics the dense, fibrotic microenvironment characteristic of late-stage PDAC [12]. Our findings revealed that G43 cells grown in softer ECM were initially more sensitive to treatment compared to G46 cells under similar conditions (Figure 5A). Interestingly, when G43 cells were cultured for a longer duration in stiffer ECM, they exhibited increased sensitivity to treatment. Conversely, G46 cells showed no significant differences in treatment sensitivity between soft and stiff ECM (Figure 5B). The mechanism underlying this differential sensitivity in G43 cells remains unknown but suggests that ECM stiffness may influence cellular responses to treatment, potentially through mechanotransduction pathways that modulate drug sensitivity.

To evaluate the impact of AZD 1775 on Gemcitabine 8C efficacy in pancreatic ductal adenocarcinoma (PDAC) organoids, G43 and G46 cells were embedded in soft (3A) or stiff (6A) ECM-mimicking matrices and treated following 48-or 168-hour organoid formation. Viability was assessed using the CellTiter-Glo 3D assay and Live/Dead staining. At the 48-hour time point, combination treatment with Gemcitabine 8C + AZD 1775 led to decreased ATP levels in both cell lines, particularly in stiff ECM, while Live/Dead staining showed more viable cells in soft ECM under combination therapy. Notably, at the 168-hour time point, a divergence emerged between assays. While CellTiter-Glo luminescence indicated increased viability—especially in stiff ECM with Gemcitabine 8C or the combination—the Live/Dead assay revealed increased cell death under the same conditions. This discrepancy suggests that ATP measurements may be confounded by metabolic contributions unrelated to viability. Specifically, the Gemcitabine 8C prodrug contains an octanoic acid moiety that can be cleaved by carboxylesterases (CES1/CES2), releasing free octanoate. As supported by recent findings, octanoate can enter the tricarboxylic acid (TCA) cycle and fuel mitochondrial ATP production, potentially inflating luminescence signals independent of actual cell survival [20]. These results highlight the importance of using complementary viability assays and considering metabolic artifacts when interpreting prodrug responses in 3D cancer models.

Organoid models derived from G43 and G46 pancreatic cancer cells exhibited differential responses to Gemcitabine 8C and AZD 1775 treatment depending on ECM stiffness and duration of pre-formation. After 48 hours of organoid formation, combinational treatment with Gemcitabine 8C + AZD 1775 increased cell viability in soft ECM (3A) but reduced viability in stiff ECM (6A), as shown by Live/Dead staining. Acridine orange staining revealed an increase in double-stranded DNA (DS-DNA) signal with AZD 1775, suggesting cell cycle arrest or inhibition of DNA processing. In contrast, Gemcitabine 8C alone led to increased single-stranded DNA (SS-DNA), consistent with replication stress or DNA damage. After extended 168-hour organoid formation, both G43 and G46 models showed reduced viability across all treatment conditions, with more pronounced cell death in both soft and stiff ECM, as indicated by a reddish merged Live/Dead signal. Acridine orange staining at this later time point revealed a shift toward more single-stranded DNA and acidic compartment accumulation, particularly under combination treatment, as evidenced by yellow-orange fluorescence, suggesting enhanced DNA damage or apoptosis. Together, these results indicate that ECM stiffness and the timing of treatment critically influence therapeutic response and cell fate in PDAC organoid models.

The findings from this study underscore the potential of using PDO models to develop personalized treatment strategies for PDAC patients. By recapitulating the unique TME and heterogeneity observed in individual patient tumors, PDOs allow for drug screening in a physiologically relevant context. The differential responses observed between G43 and G46 cells in distinct ECM environments highlight the importance of considering tumor heterogeneity and microenvironmental factors when designing treatment regimens. Future studies should aim to elucidate the molecular mechanisms underlying the differences in treatment sensitivity observed between these cell types, with a particular focus on cytoskeletal dynamics, oxidative stress responses, and mechanotransduction pathways. Based on our experimental findings, we propose a working model for the mechanism of action of Gemcitabine 8C and AZD 1775 (Fig. X). While the metabolic processing of Gemcitabine 8C remains to be fully characterized, we hypothesize that cleavage of the octanoic acid moiety may contribute to anaplerotic fueling of the TCA cycle, potentially explaining the observed ATP increases despite decreased cell viability. This schematic integrates established pathways of gemcitabine metabolism and AZD 1775-mediated WEE1 inhibition, along with our proposed mechanism, providing a framework for future investigation.

## Data Availability Statement

Data are available on request.

